# Safe Blues: A Method for Estimation and Control in the Fight Against COVID-19

**DOI:** 10.1101/2020.05.04.20090258

**Authors:** Raj Dandekar, Shane G. Henderson, Marijn Jansen, Sarat Moka, Yoni Nazarathy, Christopher Rackauckas, Peter G. Taylor, Aapeli Vuorinen

## Abstract

How do fine modifications to social distancing measures really affect COVID-19 spread? A major problem for health authorities is that we do not know.

In an imaginary world, we might develop a harmless biological virus that spreads just like COVID-19, but is traceable via a cheap and reliable diagnosis. By introducing such an imaginary virus into the population and observing how it spreads, we would have a way of learning about COVID-19 because the benign virus would respond to population behaviour and social distancing measures in a similar manner. Such a benign biological virus does not exist. Instead, we propose a safe and privacy-preserving digital alternative.

Our solution is to mimic the benign virus by passing virtual tokens between electronic devices when they move into close proximity. As Bluetooth transmission is the most likely method used for such inter-device communication, and as our suggested “virtual viruses” do not harm individuals’ software or intrude on privacy, we call these *Safe Blues*.

In contrast to many app-based methods that inform individuals or governments about actual COVID-19 patients or hazards, Safe Blues does not provide information about individuals’ locations or contacts. Hence the privacy concerns associated with Safe Blues are much lower than other methods. However, from the point of view of data collection, Safe Blues has two major advantages:

- Data about the spread of Safe Blues is uploaded to a central server in real time, which can give authorities a more up-to-date picture in comparison to actual COVID-19 data, which is only available retrospectively.
- Sampling of Safe Blues data is not biased by being applied only to people who have shown symptoms or who have come into contact with known positive cases.

These features mean that there would be real statistical value in introducing Safe Blues. In the medium term and end game of COVID-19, information from Safe Blues could aid health authorities to make informed decisions with respect to social distancing and other measures.

In this paper we outline the general principles of Safe Blues and we illustrate how Safe Blues data together with neural networks may be used to infer characteristics of the progress of the COVID-19 pandemic in real time. Further information is on the Safe Blues website: https://safeblues.org/.

## 1 Introduction

There is a dire need for timely information about the spread of the COVID-19 virus. Recently, the idea of using contact-tracing mobile device apps to help in this endeavour has received a considerable amount of attention. See, for example, [1, 2, 3, 4].

There are essentially two classes of information that are provided by such apps:

- information about the actual people that an infected person has met, and
- data that can form a basis for statistical inference and control of the epidemic.

All the contact-tracing apps that we are aware of seem to have collection of the first type of information as their primary purpose. This is not surprising. It is clearly of paramount clinical importance to identify infected people, both for treatment purposes and to prevent them from spreading COVID-19. However, as the epidemic progresses and the conversation turns to the best way of relaxing government controls, it is also very important for decision makers to understand how the epidemic behaves in the whole population. At the moment data exists only about individuals who have come to the attention of authorities because they have been infected or there has been some reason to think that they are at a high risk of infection.

In this paper, we propose a framework that is aimed exclusively at collecting the second type of information mentioned above. Compared to other frameworks, it has the advantages of

- providing population wide aggregated data in real time,
- tracking the way that the epidemic might be progressing in parts of the population that do not come to the attention of authorities, and
- being less intrusive from a privacy point of view.

In general, the spread of a virus depends on both its biological properties and the behavioural properties of the population. Biological properties of COVID-19 have been studied since the start of the outbreak, see, for example, [5]. On the other hand, population behaviour is changing rapidly due to unprecedented social distancing measures and is hard to observe and to predict. As a consequence, achieving tight real-time estimates of time-varying parameters such as *R*_eff_ (*t*), the expected number of individuals infected by an infectious person [6, p. 7], is a difficult task.

In an imaginary world, we might develop a harmless biological virus that spreads just like COVID-19, but is traceable via a cheap and reliable diagnosis. By spreading such an imaginary virus throughout the population, the spread of COVID-19 could be easily estimated because the benign virus would respond to population behaviour and social distancing measures in a similar manner. Such a benign biological virus does not exist. Instead, we propose a safe and privacy-preserving digital alternative that we call *Safe Blues*.

The Safe Blues method would use Bluetooth signals similarly to the suite of existing and emerging contact tracing frameworks Proximity [4], Blue Trace [1] and the Privacy-Preserving Contact Tracing framework currently being developed by Apple and Google [2]. However, in contrast to these frameworks, Safe Blues does not record information about individuals and their interactions. Instead, it will help understand population wide dynamics in a privacy-preserving manner.

The Safe Blues idea is that mobile devices mimic virus spread via the safe exchange of Bluetooth signals. Then, aggregated counts are reported to a server without recording private information. By periodically creating various strands of Safe Blues and repeatedly spreading them through the (mobile device) population, our analysis of the signals will help to obtain aggregate estimates of population contact. The result will be a real-time estimate of the effect of any social distancing rules that are put in place. Further, when retrospective information about COVID-19 case numbers becomes available, it can be combined with Safe Blues data to train sophisticated machine-learning procedures to estimate COVID-19 infection numbers in real-time.

As an example, in Figure 1 we present an illustration of the path of one fictitious epidemic where government interventions are taken to curb the spread of the disease. The figure also presents the paths of recorded Safe Blues activity. Decision makers only receive data on the real epidemic lagged by 15 days, but the Safe Blues information is received in real time. The epidemic begins with a proportion of 1.25% infectives, along with 50 different Safe Blues Strands that have been initialised with similar proportions of infectives.

**Figure 1:**
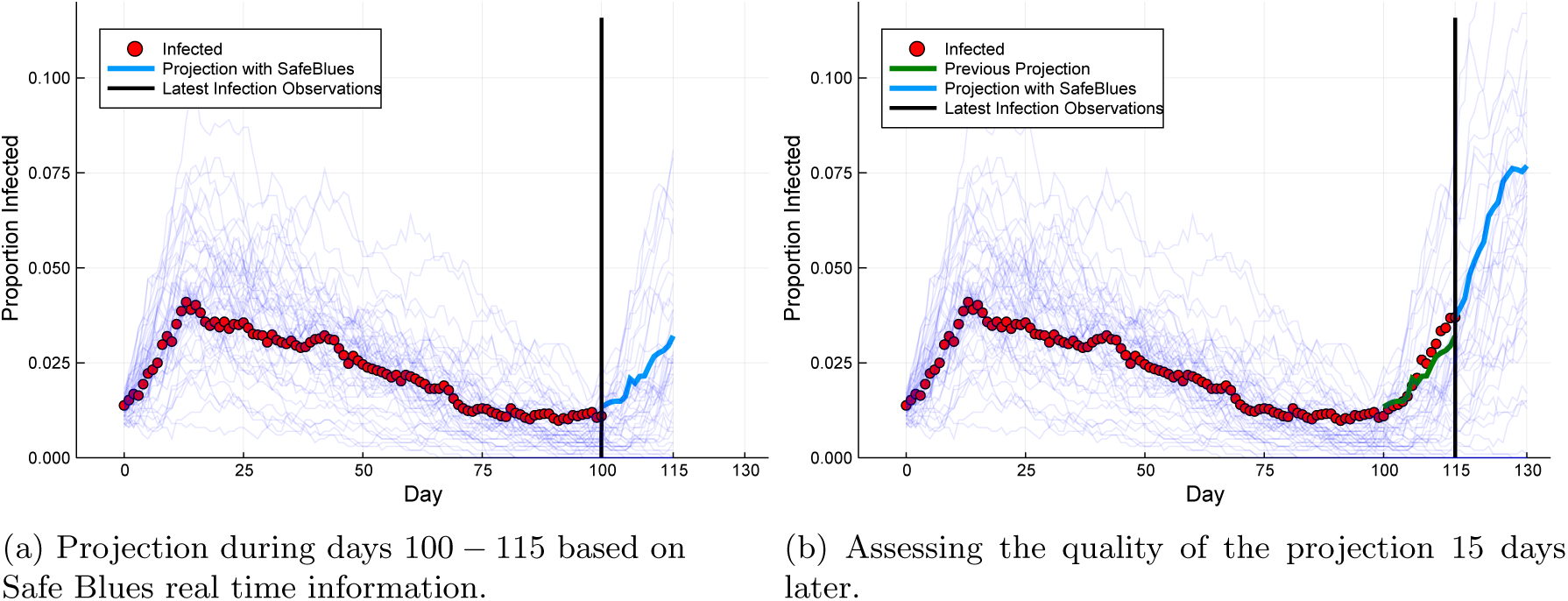
Safe Blues presents a near real-time estimate of the potential for virus spread: During days 100-115, Safe Blues activity is observed to rise and thus helps predict a rise in COVID-19 cases.

After an initially rapid spread of the epidemic, social distancing regulations are quickly tightened, and after two weeks, they are fixed to prevent the vast majority of social contact. As these rules go into effect, the proportion of infectives begins a slow but steady decline. All the while, Safe Blues Strands are being simulated on mobile devices. As a consequence, the proportion of Safe Blues infections mirrors the decline in real infectives, driven by a corresponding reduction in physical proximity between Safe Blues-enabled devices.

After 100 days since the start, and months of a promising decline in case numbers, the social distancing rules are mostly lifted. The first plot presents the view at 115 days: about two weeks past this change. At this point, only data up to day 100 is observed, while Safe Blues information is observed in real-time up until day 115. These two pieces of information have also been used to compute a live estimate of the epidemic, shown as a light blue curve in Figure 1a. This estimate shows that the epidemic is again on the rise, and that the reversal of social distancing rules may have been too early or too aggressive.

The second plot shows the situation 15 days later, after which the lagged dynamics of the epidemic have again been observed. Once more, the projection shows a strong uptick of infections. Compared to the projection computed earlier, one can see that the Safe Blues system mimics the dynamics and interaction patterns of individuals with high accuracy to yield valuable information on the real-time behaviour of the epidemic. This reflects the reality that we are observing with the COVID-19 epidemic: a long incubation period and mild symptoms at the start of infection mean that diagnoses are delayed by weeks and real-time information is almost non-existent. In this way, the Safe Blues framework may provide unique, invaluable visibility into the current state of the epidemic and a powerful tool for early detection of subsequent waves or outbreaks.

In addition to such real-time early warning predictions, Safe Blues has the potential for more. At the moment, when a government adjusts social distancing directives, it is not clear what the effect on population behaviour is and, even if adherence is immediate, time will elapse before the effect of the measures on the spread of COVID-19 becomes observable. In particular, this makes it difficult to set social distancing measures in a way that balances objectives while keeping *R*_eff_ (*t*) sufficiently low such that the epidemic does not take off again. This is an important consideration towards the end game of COVID-19. The Safe Blues idea will produce a faster feedback loop because indicators of social contact can be observed in a more timely fashion.

If Safe Blues is implemented in the early or middle stages of the COVID-19 pandemic, information obtained during that period will be beneficial for later decision making, especially when dealing with efforts to eliminate second or third waves of infections. The initial analysis of this paper indicates that Safe Blues data may be useful for future projections of the epidemic. For this, we rely on statistical machine learning methods, mixed with solid principles of epidemiological modelling. As COVID-19 progresses, the estimates will become more and more precise. A consequence is that within three to six months of deployment of Safe Blues, estimates of *R*_eff_ (*t*) for COVID-19 could be tighter than they currently are. Similarly, the effect of various forms of social distancing government directives on *R*_eff_ (*t*) will be better understood. As an end result, governments will have a better grasp of how to optimally “flatten the curve” while keeping the economy as active as possible.

### Structure of this paper

In Section 2 we describe the motivation and underlying principles of Safe Blues. We also discuss the level of privacy that Safe Blues offers (Section 2.3) and the potential for integration with current contact tracing apps (Section 2.4). In Section 3 we illustrate the power of Safe Blues for estimation, future projections of the epidemic, and control. For this we present two methods that we call *Deep Safe Blues* and *Dynamic Deep Safe Blues*. We then conclude in Section 4. Further information is also available on the Safe Blues website: https://safeblues.org/.

This paper also contains an extensive set of appendices. In Appendix A we overview the software and the protocols of communication needed for Safe Blues. In Appendix B we spell out some of the details of the machine learning methods employed for estimation and projection. In Appendix C we describe the three test bed models that were used to evaluate the potential power of Safe Blues.

## 2 Underlying principles

In the modelling of epidemics, the effective reproduction number *R*_eff_ (*t*) is the central quantity that determines how the epidemic grows or diminishes. This quantity is defined as the average number of individuals infected by each sick individual at time t. For COVID-19, early estimates indicate that, without significant control measures being in place, *R*_eff_ (*t*) lies in the range of 2 — 4, [7]. However, *R*_eff_ (*t*) depends on a combination of biological and behavioural factors. Some key biological factors include the propensity of the pathogen to infiltrate human hosts, the duration of the disease, and the susceptibility of different age groups. Some key behavioural factors include personal hygiene practices, hand shaking practices and, importantly, the proportion of time that individuals are in physical contact or close proximity. The biological factors tend to be uncontrollable and, with the exception of weather effects, may be assumed to remain constant as long as significant virus mutation does not occur. However, the behavioural factors are controllable, at least to some extent. Indeed the suites of social distancing measures imposed in over 150 countries during the first few months of 2020 are attempts to control the behavioural component of *R*_eff_ (*t*), [8]. Such social distancing measures, some of which are outlined in the impactful report [9], have been introduced to slow down the spread of COVID-19. Nevertheless, at this early stage, it is very difficult to quantify the effect that any particular social distancing measure is having on *R*_eff_ (*t*) and the dynamics of the pandemic.

Such lack of quantifiability is problematic because all models attempting to aid policy makers by projecting the course of the epidemic require an estimate of *R*_eff_ (*t*). This is often obtained by modelling that attempts to quantify the level of human to human interaction either at broad scales as in [6, 10, 11] or at finer scales as in [9, 12, 13]. A notable recent attempt to include such a quantification is described in [14] where the authors used survey sampling of the UK population to estimate that *R*_eff_ (*t*) shifted from around 2.6 prior to lockdown, to around 0.62 after lockdown, which occurred in March 2020. While impressive, such questionnaire-based surveys are difficult to execute and are not able to yield real-time estimates of *R*_eff_ (*t*). Other attempts at measuring *R*_eff_ (*t*) use up-to-date counts data such as the now famous dashboard by the CSSE at Johns Hopkins [15]. An example of such dynamic prediction is presented in the dashboard [16] which pulls counts data from [15]. However, in such cases the problem is that reported “live data” about COVID-19 is based only on confirmed tested cases and does not consider the large number of asymptomatic cases or untested cases that must exist. Better estimates become known only retrospectively, after the pandemic has progressed.

At this time (April-May 2020) a considerable number of countries still have a significant amount of infection, and many have imposed a suite of social distancing measures that together seem to be having the effect of reducing *R*_eff_ (*t*). However, it is not clear which individual measures are driving this. In the second half of 2020 and onwards, getting more precise estimates of effectiveness will be very important. Many governments will grapple with the optimal way of lifting (and at times reinstating) social distance measures as they attempt to balance economic revival with health considerations. For this, having fine-tuned live estimates of *R*_eff_ (*t*) and related quantities is of paramount importance. Safe Blues is designed to serve as a tool that can help in this arena. If it is introduced at around May-July 2020, then after a training and calibration period, by September 2020 and onwards the tool has the potential to serve as an aid to policy makers when they are considering the adjustment of social distancing directives. Figure 2 presents a simple illustration of such a timeline.

**Figure 2:**
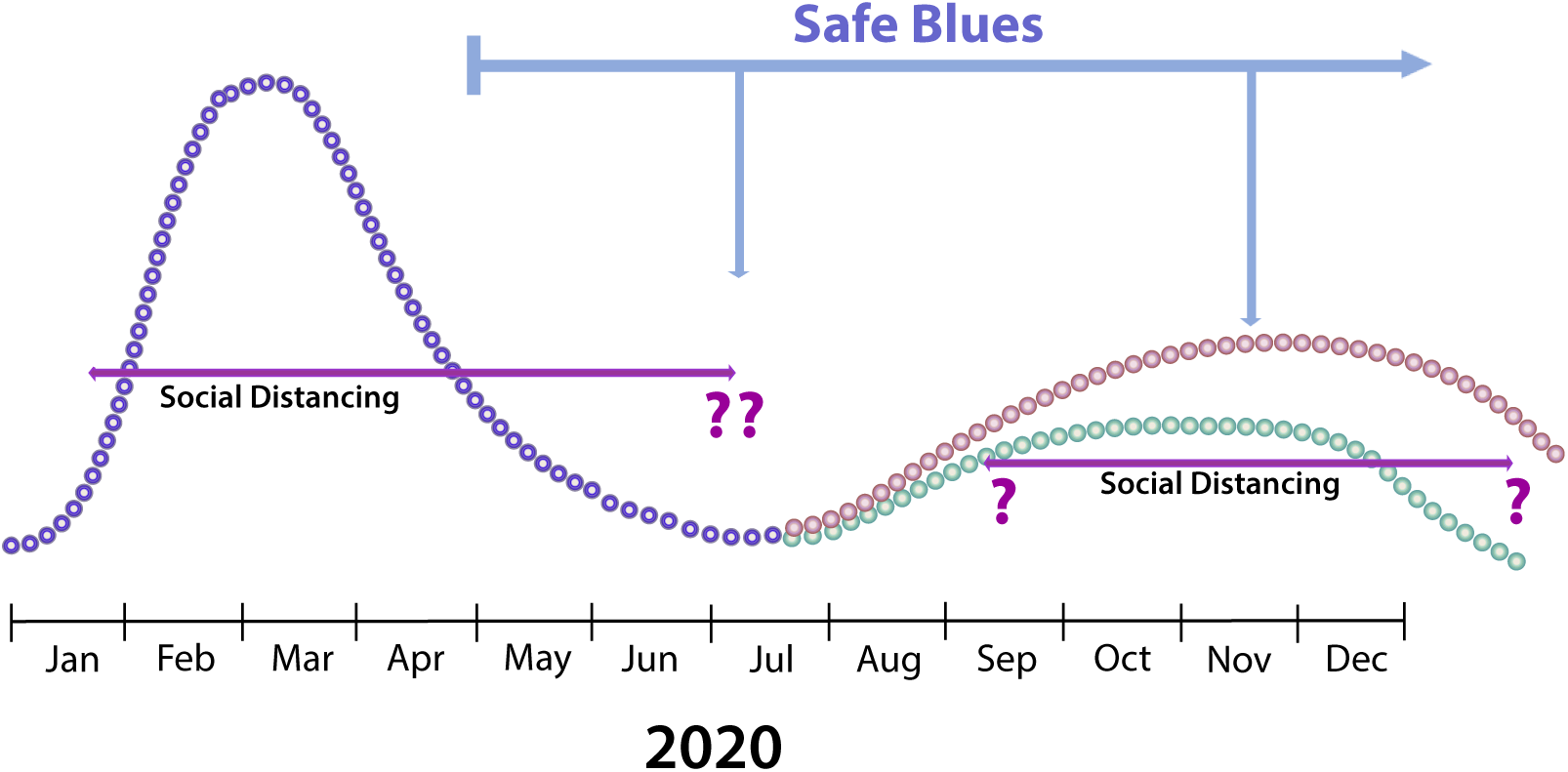
An illustrative timeline of the epidemic in a particular region. Introduction of Safe Blues at around May-July may imply that by September meaningful insights about the epidemic can be obtained. This can help to inform social distancing policy in the second half of 2020.

### 2.1 How it Works

The key idea of Safe Blues is to obtain real-time estimates of gross population engagement dynamics in a safe and privacy-preserving manner. In real time, Safe Blues data can be processed to yield estimates of COVID-19 *R*_eff_ (*t*) as well as other parameters that can be used to inform future epidemiological models.

The system works by having personal mobile devices take part in an ongoing *safe real-time virus spread simulation* where, by means of Bluetooth signals, the time that individuals spend in close proximity is a key driving factor. This is done in a way that does not compromise individual privacy, does not cause any risk to human health, and does not introduce any risk to individual software or hardware. See Figure 3 for a schematic illustration of the Safe Blues system.

**Figure 3:**
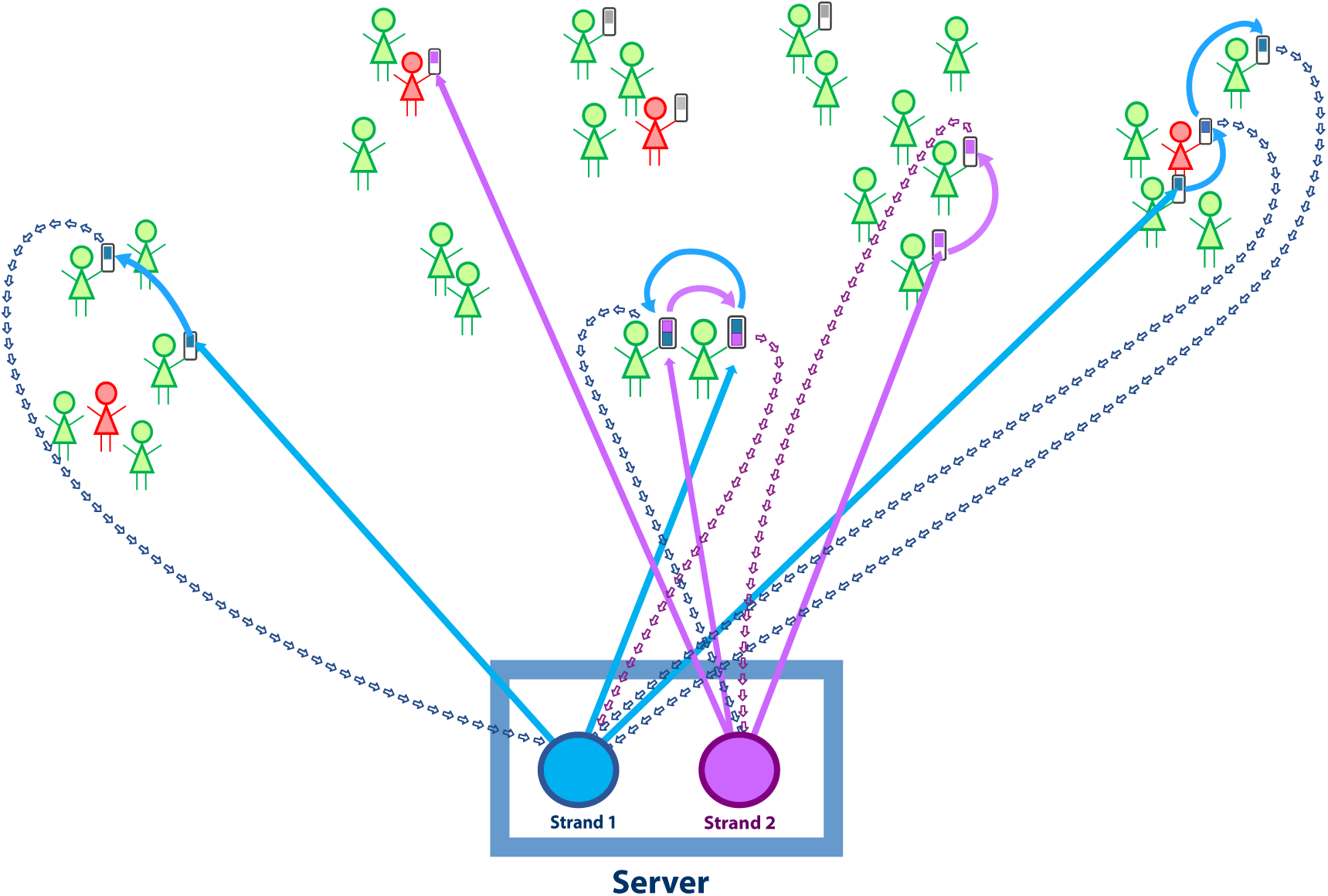
Individuals of the population with Safe Blues enabled devices take part in spreading Safe Blues Strands. Marked individuals support the Safe Blues system by carrying devices with Safe Blues software. COVID-19 infected individuals are in red and others are in green. The Safe Blues system operates independently of the COVID-19 status of individuals.

A *Safe Blue Strand* is a virtual token that circulates and replicates between the mobile devices of individuals using dynamics designed to reflect the transmission of an actual biological virus but without any threat to safety, software, or privacy. Similar to a biological virus, the harmless Strand is counted as “active” for a finite duration of time in each mobile device that is “infected”. During that time, if the mobile device is in close proximity to another device, there is a chance for the Strand to “spread” to the neighbouring device. Similarly, if the mobile device is in relative isolation, the Strand is not likely to spread and the mobile device will eventually “recover”. Further, as with actual biological viruses, Strands can have an incubation period during which they are not infective.

By allowing multiple types of Strands, or multiple Strands, to “infect” the mobile device population, the “epidemics” of the Strands respond to social mobility and social distancing measures in a similar (but not identical) way to COVID-19 response. However, in contrast to COVID-19, the number of devices “infected” by Strands can be measured in real time.

The Safe Blues system will periodically “inject” such Strands into the mobile host population and obtain real-time counts of the number of “infected” hosts for each Strand. While the population dynamics of each Strand will not directly resemble the dynamics of COVID-19, the underlying driving mechanism of close physical social interaction will be shared by both the real biological virus and the harmless Strands. Hence, we expect the course of Strand “epidemics” to be coupled with the course of the COVID-19 epidemic, especially under varying social distance measures.

The mechanism of communication between devices is short-range Bluetooth. This is similar to the communication protocol used by many emerging contact tracing apps (see Section 2.4 below). In such a setting, as individuals spend time in close proximity, the propagation of these virtual Strands has a higher tendency to succeed and spread. Conversely, as individuals maintain a higher level of social distancing, the Safe Blues Strands are less likely to spread. Unlike systems that use contact tracing apps, the Safe Blues system is oblivious to the actual health status of specific individuals. For example, in Figure 3, some individuals are infected by COVID-19 (red) while others are not (green). However Safe Blues is not aware of and does not need this private information. Similarly, some individuals participate in Safe Blues (as signified by a grey “mobile device”) and others do not. Clearly some level of population participation is required, but Safe Blues does not require all individuals to participate.

Figure 4 illustrates the outcome from a simulation of the COVID-19 epidemic in parallel to multiple Strand trajectories. Specific details about our simulation test beds are given in Appendix C. The key point illustrated in Figure 4 is that multiple Strands can co-exist in parallel to the COVID-19 epidemic and that they are influenced by social interaction and social distancing in a similar way to COVID-19.

**Figure 4:**
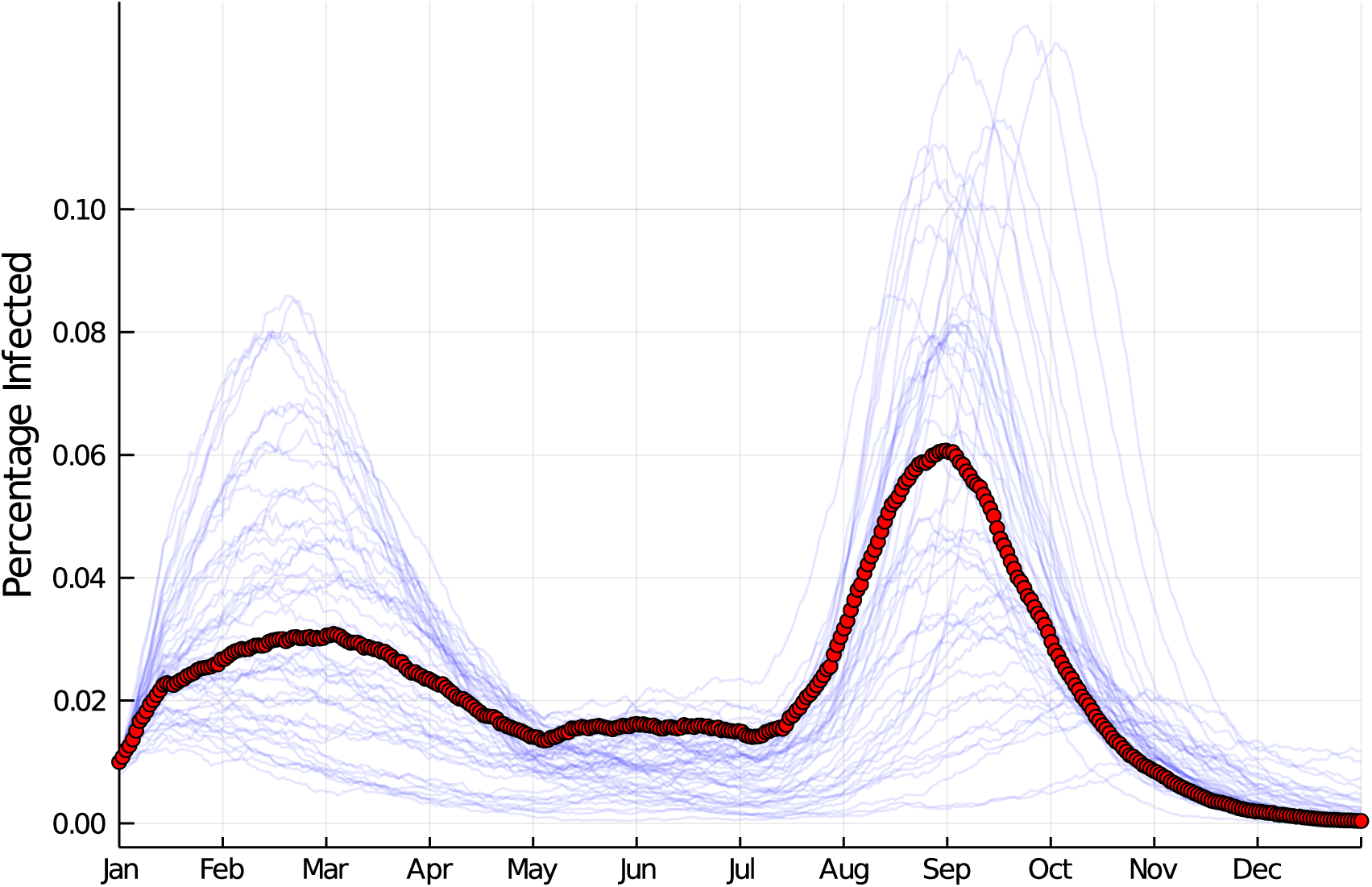
A potential course of the epidemic (number infected) with associated Safe Blues Strands. Social distancing measures modify the course of the epidemic and in the process influence Safe Blues.

### 2.2 Further Details

Each Strand is uniquely identified by an integer *s*. The Strand has associated start time *t*_start_ at which the server *seeds* the Strand with the participating mobile devices. As this occurs, each device independently chooses whether to get “infected” by the strand, with probability *p*_0_, or ignores the seeding otherwise. Thus devices generate random outcomes that affect the (Safe Blues Strand) epidemic. Hence at *t*_start_, if there are *N^B^* mobile devices with Safe Blues enabled, there will be approximately *p*_0_ × *N^B^* devices infected with the specific Strand.

As an individual together with their Safe Blues-enabled device physically nears another individual with such a device, Bluetooth is used to transmit active Strands from the infected individual to the other. This communication involves a short-range distance measurement denoted by *D*. The duration of the physical proximity, denoted by *τ*, is also estimated. Then the probability of infection is given by a function *I_s_*(*D*,τ), specific to Strand *s*. Although no Strand will behave exactly like COVID-19, statistical analysis will later reveal which choices of the function *I_s_*(*D*,τ) yield better fits to COVID-19 data.

Once a device is infected by the Strand, it enters a random *incubation period*. During that period the device “carries” the Strand but cannot infect other devices. Once the incubation period is complete, the device becomes “infective” and starts to infect other devices that are in close proximity. This occurs until the device has “recovered”.

Appendix A provides further details on the suggested system architecture from a software engineering point of view.

#### On the Penetration Proportion and Strand Parameters

We use *N* to denote the number of individuals in a population. In determining the parameters of a Strand, the *penetration proportion η* = *N^B^*/*N* plays a key role. Assume that at a given point in time, COVID-19 has an estimated infection rate of *β* and removal rate of *γ*. We can then set the infection rate and recovery rate of strand *s* to roughly follow

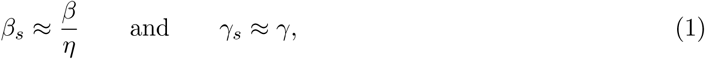

where the approximation is due to the lack of exact knowledge about *γ* and *β*, as well as due to the desire for some heterogeneity between strands. That is, we wish for the strands to have different attributes, yet they should roughly reflect the behaviour of COVID-19. The choices of the parameters *β_s_* and *γ_s_* can then be incorporated into actual operational strand parameters such as *I_s_*(·, ·) and the infection period distribution, with details appearing in Table 2. For the incubation period, we may want to set the strand parameters to be lower than typical COVID-19 parameters, for quicker response time in measurements.

The motivation for Equation (1) comes from basic epidemiological considerations appearing in SIR models. As an example, consider the difference equations associated with Model I (see Appendix C). In this case, if one decreases the population size by a factor of *η*^-1^, then achieving similar epidemic behaviour (on the smaller population) can be achieved by setting *β_s_* and *γ_s_* as in Equation (1).

As for the needed penetration level of Safe Blues, the larger a population is the smaller the penetration proportion *η* can be while still generating useful data. Numerical experiments suggest that in a relatively small population of *N* = 100, 000 individuals an *η* between 0.1 and 0.2 already gives high-quality data. We hypothesise that with N in the order of millions a penetration level *η* between 0.05 and 0.1 may be sufficient for a successful estimation using Safe Blues. It is important to note that the required penetration level for Safe Blues is much lower than for contact-tracing apps to be successful.

### 2.3 Privacy with Safe Blues

Implicit in the design of Safe Blues is an important privacy feature: no individual interaction information or any other private information is shared between devices or between a device and the database. The entire protocol runs without associating long-term identifiers with users and no user can ever know the identity of any other user. In fact, users need not share their location, name, number, identity, health or infection status, movement patterns, or any other type of personal or identifying information with the app. Similarly, the devices do not share anything else between each other than the Strands with which they are currently infected. A specification of the information transmitted between Safe Blues devices and the server is in Appendix A.

This is in contrast to contact-tracing apps that raise more serious concerns about personal privacy, even when engineered using novel privacy-protecting methods. Fundamentally, this is because the goal of any contact-tracing app is to observe relationships between individual people through their interactions, whereas the goal of Safe Blues is to collect only aggregate simulated epidemic signals. Hence, Safe Blues can be implemented in a way that preserves privacy to a greater extent than any currently-proposed or implemented contact-tracing solution. See for example [1, 4, 17, 18]. Thus a proper implementation of Safe Blues can provide stronger privacy guarantees than apps attempting to mitigate COVID-19 through contact-tracing methods.

Nonetheless, there are some issues that need to be addressed. One is the case of an adversary choosing a rare Strand and infecting a user with that Strand, then tracking the spread of that particular Strand to attempt to track the user. In extreme cases where only very few users are affected by a Strand, a third party may be able to achieve some sort of de-identification. This, however, can be mostly subverted by making sure that the seeding probability of each Strand is sufficiently large to make Strands common. Note also that to transmit a rare Strand, the adversary would need to be within several meters of the target for an extended period of time. The fact that the seeding has to occur on each device makes sure that an impostor of the Database cannot perform a similar attack.

In large deployments, governments and health authorities implementing Safe Blues may wish to gain fine-tuned information regarding the geographic patterns and potential spread of the virus. For this, they could add a local identifier in the app. However, such an identifier should refer only to a general region such as a country, state, or a major city. This is also approximately the same level of granularity at which users can be trivially tracked through their internet address, and a myriad of other well-established techniques.

Finally, there may be concern that each device must connect back to a central Database each day. This is a valid concern, but must be understood in the context of modern apps, where most users already accept regular connection back to servers. This is a widespread practice in almost every app, for various reasons including automatic updates and bug reporting. With any such connection there is some privacy leakage through internet addresses and other identifiers of traffic patterns. However, with Safe Blues, the Hosts do not share any long-term identifiers with the server, and there is no way for a server to tag a user across multiple database pushes. This makes it impossible to perform tracking or de-identification beyond that provided by, for example, knowledge of any website that a user visits.

### 2.4 Potential for Integration with Existing and Emerging Apps

To the best of our knowledge, the Safe Blues system significantly differs from all existing and emerging apps dealing with COVID-19. Contact-tracing apps are concerned with individuals and interactions, whereas the Safe Blues system estimates general population dynamics. Nevertheless, in terms of the software and hardware infrastructure, there is room to embed the Safe Blue protocol within contact-tracing apps in a simple manner. Alternatively, one may consider implementing a stand-alone Safe Blues app, in which case “marketing” the app through a trusted organisation is essential to gain a user base, similar to any other app of this kind.

At the time of writing, multiple countries and organisations have already implemented contact-tracing apps using Bluetooth, with several additional countries rapidly developing their own solutions. Relevant research papers include [3, 4], as well as the recently published white paper associated with the Singaporean app TraceTogether [1]. Several of the organisations developing these apps have also made the source code available.

We examined the source code of some of the apps and observed that implementing the Safe Blues protocol within them would generally be a straightforward extension to the existing work. In Table 1 we list current contact-tracing apps that use Bluetooth where, if not stated otherwise, an app is available for both Android and iOS. At the moment, many of these apps require running in the foreground. However, Apple and Google are rapidly developing APIs (Application Program Interfaces) for contact tracing, [2]. These APIs will allow government-supported apps to use Bluetooth communication in the background.

**Table 1:**
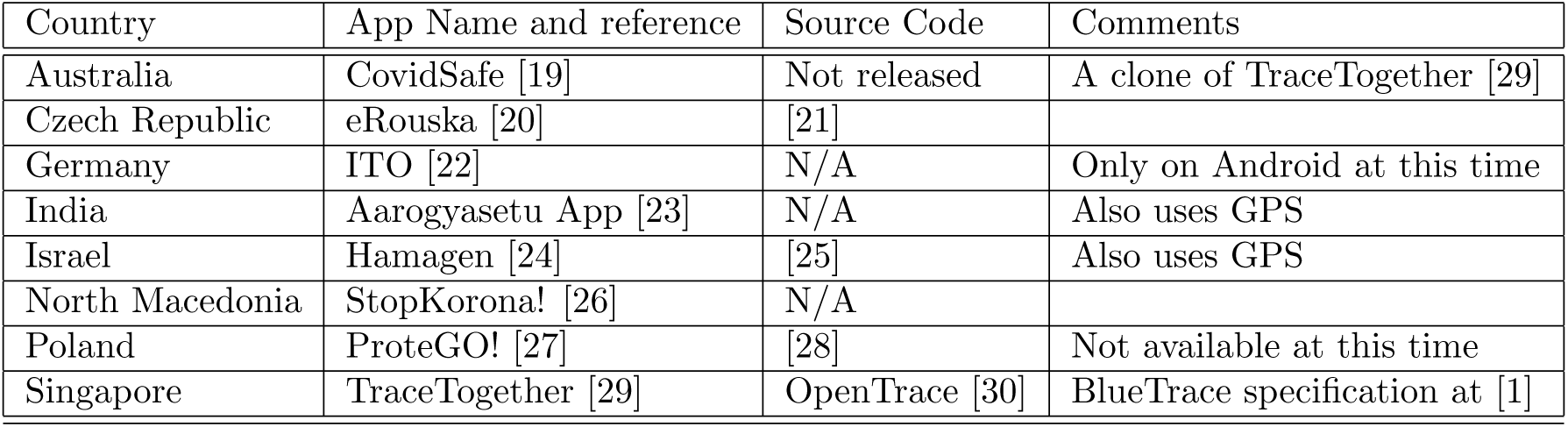
Current contact-tracing apps that use Bluetooth.

## 3 Using Safe Blues for Accurate COVID-19 Projections

While contact tracing apps can allow public health researchers to identify specific individuals who might possibly be infected, the focus on the population of known infected individuals leads to a heavily-biased sample. This does not aid in estimating the overall spread of the infection. We now illustrate how our alternative approach via Safe Blues is able to give sufficient information for estimating the real-time spread of the disease and allows for estimating the potential effects of social distancing measures.

Our methodology relies on tools from machine learning, including Deep Neural Networks, see for example [31] for an introduction. We also rely on methods for fitting UODEs (Universal Ordinary Differential Equations) as in [32]. While these are advanced mathematical and statistical tools, using our methodology is simple from a user perspective.

The basic setup follows the paradigm presented in Figure 1 where information of COVID-19 is available up to a certain point, after which only Safe Blues information is available, typically with a delay in the order of two weeks. This represents the fact that COVID-19 information is not present in real time, in contrast to Safe Blues. The relative magnitude of historic social distancing measures also is available as input for projections (for example “full lock down”, “partial lock down”, etc.).

Our analysis aims to illustrate the predictive power that can be gained by utilising Safe Blues information. For this we created three models that aim to mimic the true spread of an epidemic while taking into account the interaction between individuals via mobility or other means. A detailed description of the models is in Appendix C. The purpose of these models is not to create a detailed representation of COVID-19 spread, but is rather to supply test beds for our projection methodology. Importantly, the projection methods that we present here do not rely on the specific form of these models. In fact, these specifics are not needed for understanding how Safe Blues can be used for projection and estimation. All that is important is that each model simulates a COVID-19 epidemic, subject to time-varying levels of social distancing, and captures the Safe Blues dynamics, also affected by the same social distancing measures. We label the models as Model I, Model II, and Model III, and use a single simulation run for each model. The details of the parameters used for the simulation runs are also in Appendix C.

Our goal is to develop methods for using Safe Blues information for a variety of tasks. These include:

1. Early warning of a rise towards a “second wave”.
2. Understanding the effect of various social distancing regimes on *R*_eff_ (*t*).
3. Designing optimal control policies for fine tuning social distancing measures towards the end game of COVID-19.
4. Projecting the course of the epidemic in the medium and long run.
5. Estimating the proportion of asymptomatic carriers of COVID-19.
6. Computing uncertainty bounds for projections.
7. Optimally choosing parameters and timing for newly created Safe Blues Strands.

We now demonstrate the potential power of Safe Blues for (1) and (2) above. (1) is achieved using a neural network model that we call *Deep Safe Blues*. (2) is achieved using a universally fitted ODE model that we call *Dynamic Deep Safe Blues*. We overview the results of these predictive methods and leave analysis of methodology dealing with (3)-(7) for future work.

### 3.1 Deep Safe Blues: Early Detection of a Second Wave

We created a deep neural network model that can be trained in real time based on the ensemble of Safe Blues strands and historical COVID-19 information. We call this *Deep Safe Blues*. It is able to accurately detect the start of a trend towards a second peak in the number of infected individuals a significant time before such data is available. Figure 5 demonstrates the application of Deep Safe Blues for simulation traces from Models I, II, and III. Importantly, the same neural network architecture was used for all three simulation Models.

Our results demonstrate the strength of Safe Blues for early detection of a second wave and showcase that the auxiliary information provided by Safe Blues Strands can be valuable for detecting the start of a second peak. This can enable public health officials to respond during the essential early period before infection estimates can be updated.

**Figure 5:**
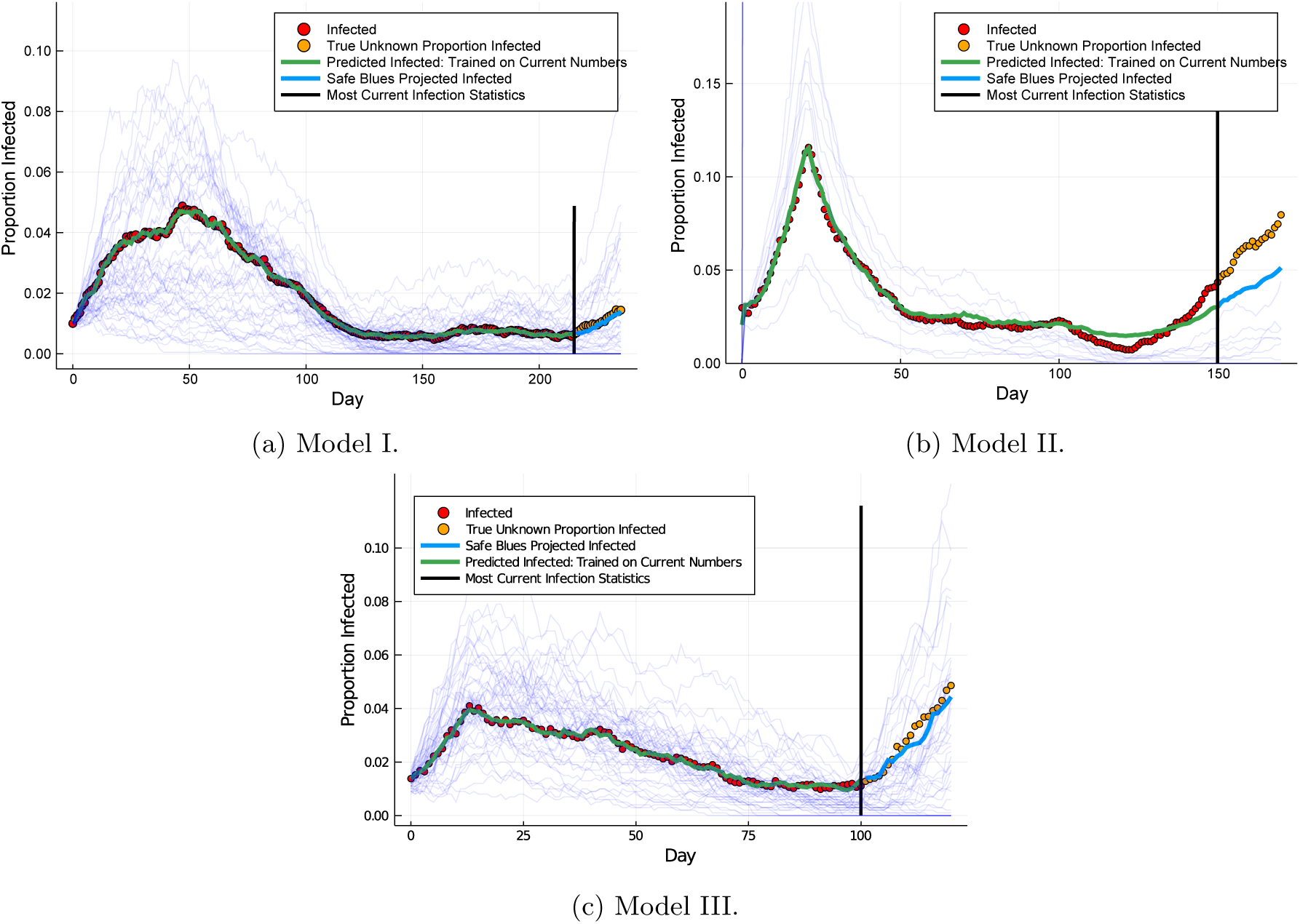
*Deep Safe Blues:* Safe Blues detection of a second wave applied to data generated from Models I, II, and III. The proportion of infected individuals is only known until the vertical black lines. After that point, only Safe Blues information is available. Nevertheless, Deep Safe Blues (trained up to the black line) is able to accurately predict a second wave of COVID-19 attack.

A full specification of the fitting methodology is provided in Appendix B.

### 3.2 Dynamic Deep Safe Blues: Policy Projection

In addition to being a tool for estimating the current number of infected individuals before such data is available, Safe Blues can also help estimate the potential effect of policy decisions. For this we developed *Dynamic Deep Safe Blues* which is a tool for projecting *R*_eff_ as a function of future levels of social distancing. Figure 6 demonstrates data-driven projections of *R*_eff_ under various policy levels. Such results can be used as input to decision makers for helping to determine the levels necessary to contain the outbreak and ensure that exponential growth into a second peak does not occur.

**Figure 6:**
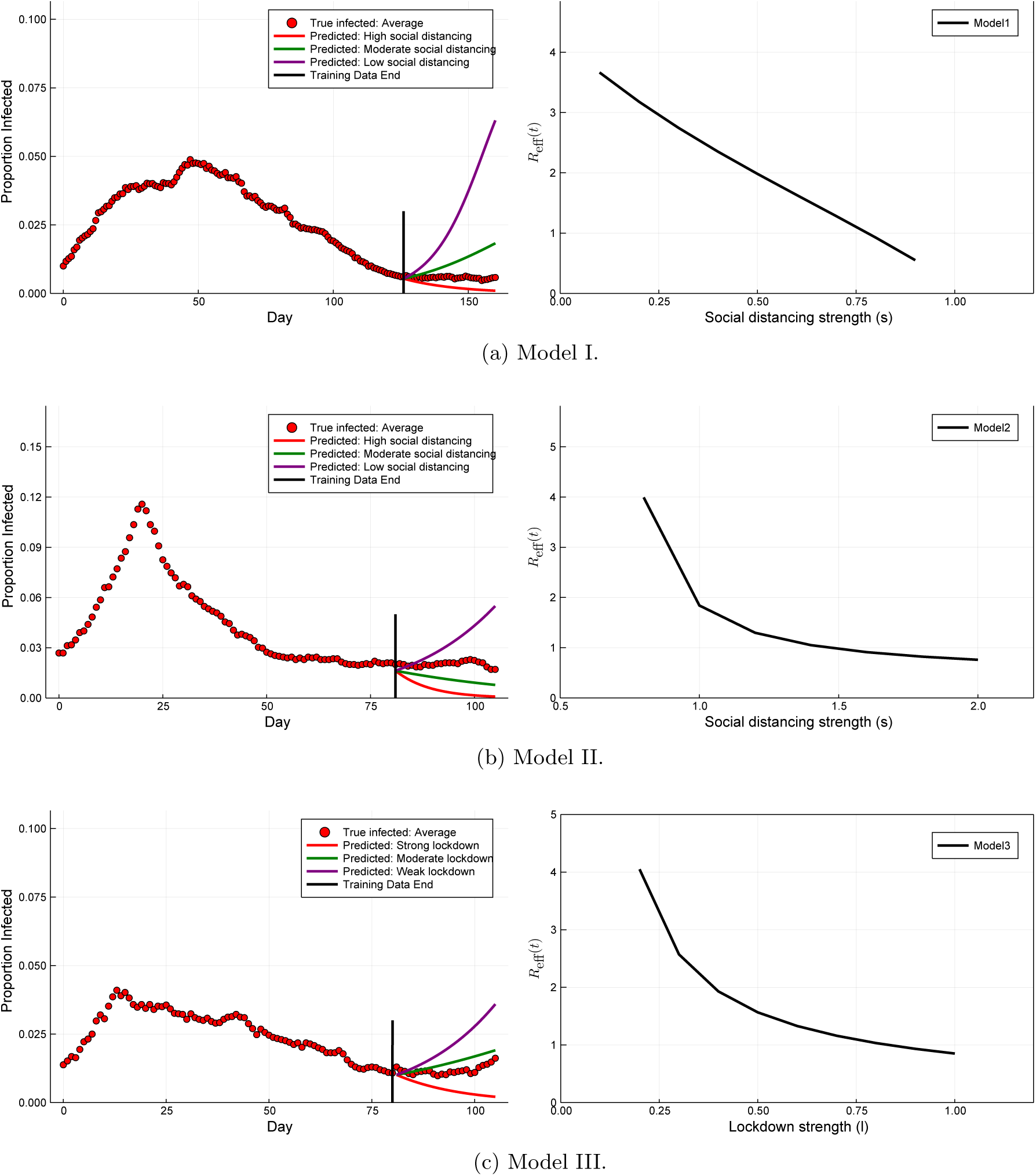
*Dynamic Deep Safe Blues:* Demonstration of policy projection and refinement on Models I, II and III. For each of these models, we predict the effect of the social distancing strength on *R*_eff_ (t_0_) where *t_0_* is the end of the training period (figures on the right). We also demonstrate potential near future trajectories as a function of policy decision (figures on the left).

The implementation of Dynamic Deep Safe Blues is based on the training of a UODE [32] using the extra information provided by the Safe Blues Strands. This approach mixes neural networks into epidemiological models in order to directly learn how policy decisions effect the spread of Safe Blues and the actual infection. Together, this allows for utilising the hidden information obtained from the simulated strands to quantify the effectiveness of social distancing approaches and determine the policies required to prevent further disease outbreak.

## 4 Concluding Remarks

This paper has presented a framework and method that can aid in estimation and control in epidemics, specifically COVID-19. To the best of our knowledge this type of framework is fundamentally different from existing solutions and other suggestions that have appeared in the literature. See for example the recent survey [33].

The machine learning principles and analysis that we used in this paper appear to be robust enough to yield immediate value from collected Safe Blues signals. We have named these *Deep Safe Blues* and *Dynamic Deep Safe Blues*. Nevertheless, there remain open questions requiring further investigation. These include designing optimal control policies for fine tuning social distancing measures, projecting the course of the epidemic in the medium and long run, estimating the proportion of asymptomatic carriers of COVID-19, computing uncertainty bounds for projections, and optimally choosing parameters and timing for newly created Safe Blues Strands.

We also mention a potential extra benefit of the Safe Blues idea. In a second or third generation of apps, one may consider presenting individual users with an up to date count of how many strands of Safe Blues their devices are infected with. One may envision that this will enable users to get a feel for the level of social distancing that they are practising and to stay socially responsible as advised by government.

We believe that Safe Blues can have significant impact in the end game of COVID-19 as it may support governments in making optimal decisions with respect to adjustments of social distancing measures. Safe Blues can be easily implemented as a layer within a current contact-tracing app, or alternatively be hosted by other COVID-19 related apps. Appendix A provides an accessible description of what such an implementation requires. Due to the critical nature of COVID-19, our team at https://safeblues.org/ is willing and able to advise and help with such implementations and with the analysis of collected Safe Blues data.

## Data Availability

Data may be obtained by contacting the corresponding author.

## Acknowledgments

We thank James McCaw for insights and advice. We thank Toshali Banerjee for help with illustrations. We are grateful for feedback and advice from Grant Sanderson (3Blue1Brown), Krzysztof Bisewski, and Itsi Weinstock. MJ and YN are supported by Australian Research Council (ARC) under grant number DP180101602. SM and PGT are supported by the Australian Research Council (ARC) Centre of Excellence for Mathematical and Statistical Frontiers (ACEMS) under grant number CE140100049.

## Appendix

### A Software and Protocol Overview

We now describe a simple and straightforward protocol for the Safe Blues system, involving “Hosts”, “Strands”, a “Database”, and a “Controller” as its basic entities. The Hosts are mobile devices carried by individuals that run Safe Blues enabled software, such as an iOS or Android mobile phone. These Hosts attempt to infect each other with Safe Blues Strands when in physical proximity, akin to how the individuals themselves might infect each other with a real virus when in physical proximity. The central Database exposes a restful API that accepts and aggregates infection reports from Hosts and provides an endpoint to download an updated list of all current Strands along with their parameters. Additionally, the Database tracks the spread of each Strand through aggregate state counts, and allows interested parties to download aggregate time-series data on the spread of Strands. Finally, the Controller is a person charged with introducing new Strands into the system in response to machine learning and forecasting needs.

Hosts do not themselves need to be uniquely identified, however an implementation may wish to include a short-lived identifier that is regenerated regularly and can be used in conjunction with information about the source of the infection reports to guard against bad actors filling the database with false reports. The notion of a locale, such as a country or state may be implemented for the purposes of more geographically fine-grained detail on Safe Blues spread. We omit such details here and present only a basic design assuming a single-locale deployment.

The main activities of the distributed system are implemented by hosts and denoted with upper-case letters, these include `PULL-STRANDS` and `PUSH-INFECTION-REPORT` for interacting with the Database; `SHARE-LIST, DISCOVER-NEIGHBORS-LIST` and `DISCOVER-NEIGHBORS-PROXIMITY` for interacting with other Hosts via Bluetooth. Finally, the activities `UPDATE-AFTER-MEET-NEIGHBOUR` and `PERIODIC-UPDATE` update the Strand states within a Host. The activities are listed in Table 5 and explained in detail later. Since a Host may be turned off, may encounter connection issues, or its timely operation may be affected in many other ways, we assume a “best effort” schedule for performing these actions. This means that if a Host misses an activity, it ought to perform it as soon as it resumes operation.

#### A.1 Strands

Each Strand, uniquely identified by a `strandID`, is defined by an immutable list of parameters presented in Table 2. For each Host, a given Strand acts as a state machine with four infection states: *SUSCEPTIBLE, INCUBATING, INFECTED*, or *REMOVED*. When a new Strand is pulled from the Database as described later, the Host is either seeded with the Strand with probability `seedingProbability` in which case that Strand starts in state *INFECTED*, or the Host is not seeded, and the Strand starts in state *SUSCEPTIBLE*. After this initial seeding, a Host may “catch” the Strand from another Host with some probability at any time between `startTime` and `endTime` given that the other Host is in close proximity and its state for that Strand is *INFECTED*. This happens at each encounter with an infected host according to a probability determined by the `infectionProbabilityMap` that depends on the mean distance and total duration of that encounter. If the Host is to catch the Strand, then that Host enters the *INCUBATING* state for a random length of time, determined by `incubationPeriodDistribution`. Note that a Host that is seeded with a Strand does not undertake this incubation period. Regardless of how a Host transitioned into the *INFECTED* state, it stays in that state for a random time determined by the `infectiousPeriodDistribution`. Finally, after being in state *INFECTED* for this amount of time, the Host transitions into *REMOVED* state and stops interacting with that Strand. The mechanics of these transitions are defined in detail in later sections.

**Table 2:**
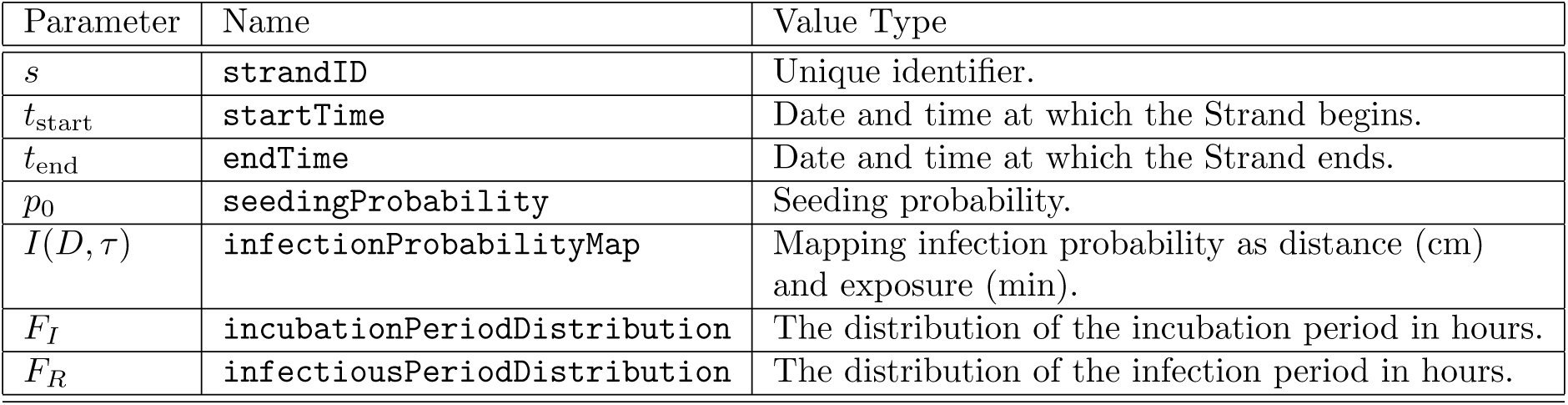
The parameters that define a single Strand.

| Parameter | Name | Value Type |
| --- | --- | --- |
| $s$ | **strandID** | Unique identifier. |
| $t_{\text{start}}$ | **startTime** | Date and time at which the Strand begins. |
| $t_{\text{end}}$ | **endTime** | Date and time at which the Strand ends. |
| $p_0$ | **seedingProbability** | Seeding probability. |
| $I(D, \tau)$ | **infectionProbabilityMap** | Mapping infection probability as distance (cm) and exposure (min). |
| $F_I$ | **incubationPeriodDistribution** | The distribution of the incubation period in hours. |
| $F_R$ | **infectiousPeriodDistribution** | The distribution of the infection period in hours. |

#### A.2 The Database

The Database exists to distribute the Strand parameters to Hosts and to track and aggregate the spread of each Strand. The Host pulls an updated list of Strands from the Database daily in an activity denoted `PULL-STRANDS` in order to discover newly introduced Strands. We describe this and other Host activities in the next subsection. The implementation should begin advertising an upcoming Strand at least one day before its `startTime`, to guarantee full propagation of its parameters prior to the start of its spread. Additionally, an implementation may wish to version Strand definitions or distribute changes to Strand definitions to reduce traffic.

The Host pushes a report of its current state for each Strand (in one batch) to the Database on a daily basis in an activity called `PUSH-INFECTION-REPORT`. In a `PUSH-INFECTION-REPORT` message, the Host sends to the Database three lists containing the set of Strands whose state is currently *INCUBATING, INFECTED*, and *REMOVED*, respectively. An implementation may wish to randomly distribute these pushes throughout the day to avoid burst traffic to its servers.

**Table 3:** The daily information pushed to the Database from each Host via the PUSH-INFECTION-REPORT activity.

| Parameter | Meaning |
| --- | --- |
| **currentIncubatingStrands** | List of current Strands with state *INCUBATING* on the Host. |
| **currentInfectedStrands** | List of current Strands with state *INFECTED* on the Host. |
| **currentRemovedStrands** | List of current Strands with state *REMOVED* on the Host. |

After collecting daily pushes from Hosts, the system aggregates the individual Host states to create `totalHostsSusceptible, totalHostsIncubating, totalHostsInfected,` and `totalHostsRemoved` for each strand. This update is done in a straightforward manner based on the collected history of state information as described in Table 3. This server side aggregation uses a best effort discipline to deal with missing values and other inconsistencies. A total estimate of the number of participating Hosts is presented via `totalHosts` (this number varies over time based on Safe Blues usage). This aggregate time series data is exposed for visualisation and data-analysis purposes in an open manner where for each date and active `strandID` for that day, the fields in Table 4 are made available.

**Table 4:** Aggregate information publically available via the Database.

| Parameter | Meaning |
| --- | --- |
| **strandID** | The unique identifier of the Strand. |
| **date** | The date. |
| **totalHosts** | An estimate of the total number of participating Hosts. |
| **totalHostsSusceptible** | The estimated number of Hosts in state *SUSCEPTIBLE* for **strandID** on **date**. |
| **totalHostsIncubating** | The number of Hosts in state *INCUBATING* for **strandID** on **date**. |
| **totalHostsInfected** | The number of Hosts in state *INFECTED* for **strandID** on **date**. |
| **totalHostsRemoved** | The number of Hosts in state *REMOVED* for **strandID** on **date**. |

#### A.3 Hosts

The Safe Blues protocol runs on Hosts and communicates over Bluetooth, either in a stand alone Safe Blues app, or possibly within other COVID-19 related apps that already utilise Bluetooth communication (see Section 2.4). The Bluetooth specification defines many roles through its own state machine to facilitate communication, of which the Advertiser and Scanner roles are of importance for this discussion. An Advertiser continually broadcasts short advertising packets on pre-defined advertising frequencies, whereas a Scanner reads those packets being broadcast along with a small set of metadata such as the Advertiser address and a Received Signal Strength Indicator (RSSI). Most full featured Bluetooth devices can perform both roles simultaneously. The Bluetooth specification [34] further allows devices to pair with each other and establish sessions. In this discussion, we assume a model of Bluetooth using Advertisers and Scanners without any session creation, though it is partially incomplete due to implementation constraints, and an actual implementation might need to use slightly different interfaces, or simulate this model via Bluetooth sessions. For a discussion of practical issues and one way of implementing this, see [1].

The list of activities carried out by a Host is described in Table 5. The remainder of this subsection outlines how these activities are carried out, together with the associated data.

The local state information stored and updated by the Host is describe in Table 6. A `tempID` is used to distinguish one Host from another in a small spatio-temporal window, allowing Hosts to correlate several Advertising packets to one Host, which in turn allows for computing the total duration of close contact. The `tempID` may be a random Bluetooth address and ought to be changed regularly, such as on each hour. The `currentInfectedStrands` is the list of Strands with state *INFECTED* for the Host. Infection may occur via “seeding” or via communication with neighbouring Hosts. For each such Strand there is an infection end time listed in `strandInfectionEnd` and once infection has ended, the Strand moves to *REMOVED* state and is removed from `currentInfectedStrands` and updated to `currentRemovedStrands` (see the `PERIODIC-UPDATE` activity description below). The `currentIncubatingStrands` is a list of Strands that the Host has acquired via interaction with neighbouring Hosts and are still in the “incubation period”. These Strands have a corresponding end time in the `strandInfectionEnd` list. The Host need not maintain a list of *SUSCEPTIBLE* Strands, as those are precisely the ones not in `currentIncubatingStrands`, `currentInfectedStrands`, or `currentRemovedStrands`.

The `PULL-STRANDS` activity in the Host discovers new Strands in the Database. When a new Strand is received from the Database, a “seeding” event occurs, whereby for each Strand pulled for the first time, the Host randomly decides whether to become infected by the strand or not based on the probability `seedingProbability`. If the outcome is to become infected, then infection begins at `startTime`. Even if such a seeding infection doesn’t take place, the parameter information about the Strand should be retained because the Host may be infected by another Host later on.

**Table 5:** Activities carried out by Hosts.

| Activity | Meaning |
| --- | --- |
| PULL-STRANDS | Update list of Strands from Database. See Table 2. |
| PUSH-INFECTION-REPORT | Report Host’s state for each Strand to the Database. See Table 3. |
| SHARE-LIST | Advertise list of Strands in state *INFECTED* to Hosts in proximity. See Table 7. |
| DISCOVER-NEIGHBORS-LIST | Receive and process a SHARE-LIST from another Host as a result of Scanning. |
| DISCOVER-NEIGHBORS-PROXIMITY | Estimate `contactDuration` and `averageDistance` of neighbor Host during Scanning. |
| UPDATE-AFTER-MEET-NEIGHBOUR | Update local Strand state after a Host leaves proximity. See Table 6. |
| PERIODIC-UPDATE | Update local state information periodically (hourly). See Table 6. |

**Table 6:** Local state information of a Host.

| Parameter | Meaning |
| --- | --- |
| `tempID` | A temporary unique identifier for the Host. |
| `currentIncubatingStrands` | A list of the Strands which are in state *INCUBATING* for the Host. |
| `currentInfectedStrands` | A list of the Strands which are in state *INFECTED* for the Host. |
| `currentRemovedStrands` | A list of the Strands which are in state *REMOVED* for the Host. |
| `strandIncubationEnd` | A list of times for which each Strand in the *INCUBATING* state will change to *INFECTED* state. |
| `strandInfectionEnd` | A list of times for which each Strand in the *INFECTED* state will change to *REMOVED* state. |

When two or more Hosts are in physical proximity, each Host uses a “best effort” method to execute `SHARE-LIST, DISCOVER-NEIGHBORS-LIST` and `DISCOVER-NEIGHBORS-PROXIMITY`. The exact manner in which these activities are executed is implementation specific and may depend on the host operating system, on the exact app in which the protocol is implemented, and on hardware considerations. One possibility is for an implementation to encode the state of each Strand in a bit field, where each Strand has a fixed position in the field specified as one of its parameters; this allows for a space-efficient transfer of Strand states and an efficient way of checking which Strands need to be updated.

The `SHARE-LIST` activity Advertises the information in Table 7 from one Host to another Host in physical proximity. On the receiving side, the `DISCOVER-NEIGHBORS-LIST` activity is executed when this information received through Scanning. One possibility is for each Host to be always be Advertising their `SHARE-LIST` and simultaneously Scanning incoming messages while in the background. The `DISCOVER-NEIGHBORS-PROXIMITY` activity supplies the receiving Host with the best effort `contactDuration` and `averageDistance` values. To aid in the distance estimation, the implementation may wish to include a transmitter power or device model identifier in the Advertising message, which could be combined with the RSSI or precomputed calibration data to get a highly accurate estimate of distance. Best effort is also used to estimate when such a physical interaction is complete. At that point the `UPDATE-AFTER-MEET-NEIGHBOUR` activity is carried out.

**Table 7:** Information shared by a Host during physical proximity.

| Parameter | Meaning |
| --- | --- |
| **tempID** | A temporary and unique identifier identifying the Host. |
| **currentInfectedStrands** | A list of the Strands with which host is currently infected with. |

The `UPDATE-AFTER-MEET-NEIGHBOUR` activity is responsible for potentially “infecting” the receiving Host with each of the Strands that the sending Host is infected with but which the receiving Host is susceptible to. For each such Strand, there is an independent random outcome where `infectionProbabilityMap` is used with the estimated `contactDuration` and `averageDistance` as inputs to determine the probability of infection. If the resulting outcome is that the receiving Host should become infected then the state of the Strand in the Host is set to *INCUBATING*. This is reflected by updating the `currentIncubatingStrands` to include the given strand. Further, the `strandIncubationEnd` is appended with an entry for that specific Strand with the value set to the current time plus a random variate randomly generated from the `incubationPeriodDistribution`. In any case the `endTime` of the Strand needs to be respected and once passed, all activity regarding the Strand expires, and the Host may remove all information about the Strand once it has pushed its final state to the Database.

The `PERIODIC-UPDATE` activity ought to run on an hourly basis on Hosts and is designed to update the Strand state for each Strand as follows:

- *INCUBATING → INFECTED*: If the current time is larger than the `strandIncubationEnd` time for a given Strand, the Strand state is changed from *INCUBATING* to state *INFECTED*.
- *INFECTED → REMOVED*: If the current time is larger than the `strandInfectionEnd` time for a given Strand, the Strand state is changed from *INFECTED* to state *REMOVED*.

As such, this activity ensures that Strands progress through the “course of the disease” on Hosts. If the Host is unable to perform a `PERIODIC-UPDATE` activity due to any reason, it should perform this activity before any other activity, as to guarantee that Hosts follow the prescribed state schedule.

### B Safe Blues Projection Methodology

This appendix describes the details used in Section 3.

#### B.1 Deep Safe Blues

We used the same Neural Network (NN) architecture for all three models. Real-time projections of estimated infected populations were generated by training a NN on the *B_t,s_*, the ensemble of Safe Blues infection strands at time *t*, to predict *I_t_*, the number of infected individuals at time *t*. The neural network *I_t_* = *NN*(*B_t,s_*) was a feed-forward neural network with two hidden layers of size 64 and tanh as the activation functions. Note that the size of the layers for Model II were reduced to prevent overfitting given the significantly reduced number of Safe Blues strands. For Model I, the data was trained on the time span *t* ϵ [0, 215]. For Model II, the data was trained on the time span *t* ϵ [0,150]. For Model III, the data was trained on the time span *t* ϵ [0,100]. Each time the ADAM optimiser from Flux.jl [35] in Julia [36] with adaptivity parameter 0.01 was used for 2, 000 iterations with a loss function being the sum squared error.

#### B.2 Dynamic Deep Safe Blues

The universal ODE [32] trained a variant of the SIR model

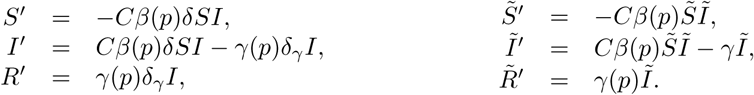

Here *C* = 0.00004 is a scaling constant, while *β*(*p*) and *γ*(*p*) are policy-dependent functions represented by neural networks. The parameters *δ* and *δ_γ_* are coupling constants used to establish a relationship between the average of the Safe Blues strands (the tilde variables) to the original infection. The neural networks had 2 hidden layers of size 16 with tanh activation functions and a final abs to ensure that the outputted values were positive without imposing bounds on the parameters. The neural networks and coupling constants were determined by minimising the Euclidian loss between the true infected and *I*, and between the mean Safe Blues infected and unfigure 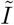. The parameters were optimised using `DiffEqFlux.jl` [37] with the `Tsit5` adaptive Runge-Kutta method from `DifferentialEquations.jl` [38]. The optimisation was done in two passes, first with the ADAM optimiser from Flux.jl [35] with adaptivity parameter 0.001.

The fitting validations for each of the models are shown in Figure 7.

**Figure 7:**
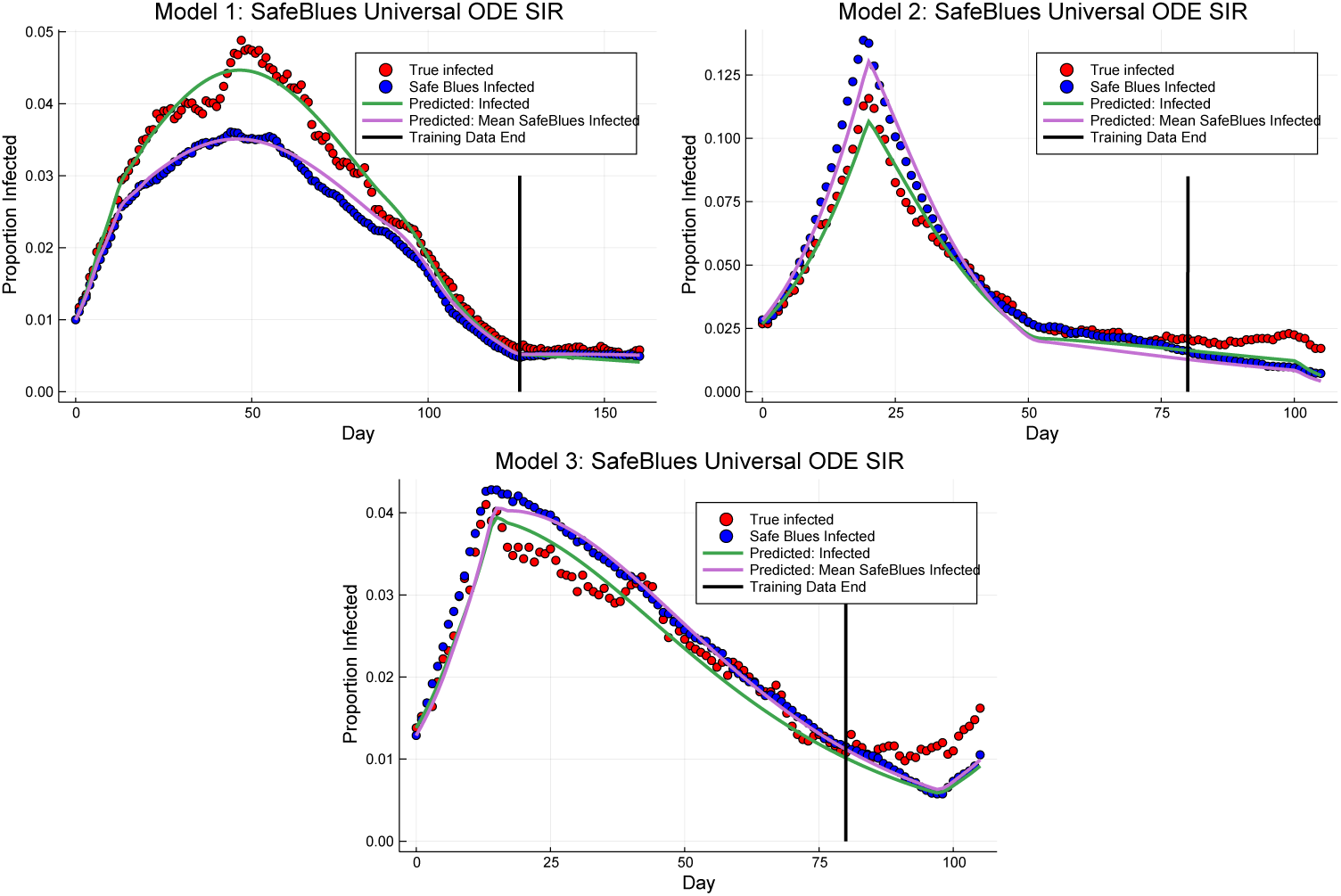
Fitting validations for the UODE models. Shown are the fits of the UODE models and their respective extrapolations.

#### B.3 Additional Remarks About Estimation

In reality, the limited availability of COVID-19 information may not be as simplistic as having a fixed delay. Nevertheless, in general the farther back we move in time, the more knowledge we have about the state of COVID-19. In practice, Safe Blues were not available at the start of the COVID-19 pandemic. See for example Figure 2 for a possible time line that incorporates Safe Blues in the fight against COVID-19. Nevertheless, for the purposes of the analysis in this section we assume availability of Safe Blues from onset. Adaptations to more realistic scenarios are possible.

The number of Safe Blues enabled users in our simulation runs was generally much smaller than is expected in reality. As a consequence, the variability of the Safe Blue infections was much higher than is expected in reality. This in turn reduces the predictive power of Deep Safe Blues and Dynamic Deep Safe Blues. However, this loss of predictive power is due to our limited simulation budget and can potentially be improved upon in an actual Safe Blues implementation.

### C The Test Bed Models

Here we outline the three different models that we use to test the effectiveness of deploying Safe Blues for projecting the spread of COVID-19. They are

**Model I**: a discrete-time stochastic SIR model.
**Model II**: a continuous-time stochastic SIR Model with migration.
**Model III**: a spatial movement model with location attraction.

Each of these models features a population comprising *N* individuals. Some of these individuals have Safe Blues-enabled devices, while others do not. At each point in time, the state of an individual registers whether they are susceptible, infected, or removed with respect to the actual virus (COVID-19). If an individual has a Safe Blues enabled-device, then the state also registers for every Safe Blues strand whether they are susceptible, infected, or removed.

The three models differ in their complexity and how they capture individual proximity. However, regardless of the model, individual proximity drives both the COVID-19 spread and the Safe Blues spread in a coupled manner, because both COVID-19 and Safe Blues only spread when individuals are in close proximity. This roughly approximates what one may expect to happen in a real scenario. Importantly, all three models allow for time-varying parameters that enforce social distancing, which in turn affects both COVID-19 spread and Safe Blues spread by changing how much time individuals spend in close proximity of each other.

Model I is a very simple and stylised model that serves as a sanity check. One of its appealing features is that converges to the well-known SIR difference equations as the population size *N* becomes large, which makes this model well suited as a first test bed. Model II incorporates several social and spatial features that are ignored in the first model. In particular, the second model has a spatial component (people have to be in the same place at the same time for virus transmission to occur) as well as a notion of social levels (people have a home where they meet a selected number of other people, a work place where they may meet a larger number of people, et cetera). Model III is a spatial model in which individuals move randomly in two-dimensional space. Its distinguishing feature is that it has a notion of centrality: although individuals move around randomly, they are biased towards visiting places that are important for them, such as their home and the supermarket. This creates a form of clustering that is not present in the first two models.

#### C.1 Model I: A Simple Stochastic SIR Model with Invitations

The deterministic discrete-time SIR epidemic model is characterised by the difference equations

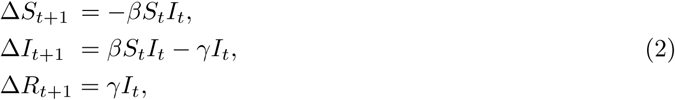

where Δ*S_t_*_+1_ = *S_t_*_+1_ - *S_t_*, and similarly for Δ*I_t_*_+1_ and Δ*R_t_*_+1_. The parameter *β* captures the rate of infection, while the parameter *γ* captures the rate of removal. Given an initial condition, the solution to these difference equations can be considered as the limit of the following simple stochastic epidemic model in discrete time.

Consider a homogeneous population of size *N*. At time *t* there are *S_t_* susceptible, *I_t_* infected, and *R_t_* removed individuals. Each individual *x* invites a fixed number *c* of randomly chosen individuals to meet. If individual *x* invites and meets individual *y*, then *x* transmits the disease to *y* with infection probability *p* = *β*/*c* if *x* has status infected and *y* has status susceptible. After all meetings have taken place, the individuals update their status for time *t* + 1. A susceptible individual *y* becomes infected if the disease has been transmitted to *y* during one of the meetings. An infected individual *x* gets removed with removal probability *γ*. The fraction of susceptible, infected, and removed individuals converges to the solution of the difference equations (2) if the population size *N* becomes large.

Including Safe Blues strands is straightforward. We simply seed the mobile devices of a number of Safe Blues users with strands. Then the spread of the Safe Blues is similar to the spreading of the actual virus. The coupling between the Safe Blues and the actual virus arises because both Safe Blues and the actual virus can be only transmitted during meetings between individuals. A schematic illustration of this process is provided in Figure 8.

**Figure 8:**
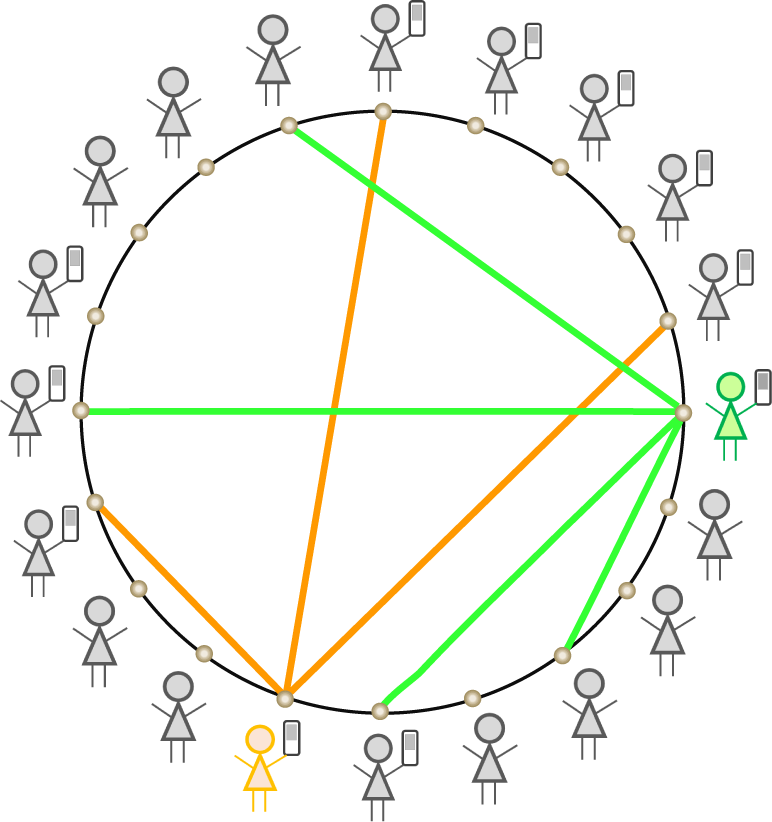
At every time point, each of the N individuals selects a random number of other individuals to invite and this implies physical proximity. In this case orange and green individuals make invitations.

To integrate social distancing in the original model, we consider the number of invitations *c* per individual as a random variable instead of a fixed number and make the mean number of invitations time-dependent. Specifically, at time *t* individual *x* invites *c_t,x_* randomly chosen people to meet, where *c_t,x_* is an independent random variable having a (truncated) Poisson distribution with mean *m_t_*. This means that the total number of meetings at day *t* is given by 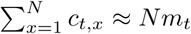. In this case the limit of the system is characterised by the difference equations (2) with *β* replaced by *β_t_* = *pm_t_*, where the infection parameter *p* is given as a model parameter.

##### Parameters used for the simulation run

We used the following model parameters to generate the data sets for the projection and policy evaluation experiments. Time consists of 366 days representing the year 2020. The population size *N* = 10^4^ and a fraction 0.2 of the population has a Safe Blues enabled device. The infection probability of the biological virus is *p* = 0.04, while the corresponding removal probability *γ* = 0.1. We introduce 50 different Safe Blues strands, with strand s having infection probability *p_s_* given as a point in the equidistant grid from 0.75(*p*/0.2) to 1.25(*p*/0.2). The removal probability *γ_s_* for strand *s* is set equal to *γ*. The epidemics for the true virus and the 50 Safe Blues strands start at day 1 with a fraction 0.01 of infected individuals in the relevant (sub)population, while the others are susceptible. The number of invitations at day *t* for individual *x* follows a (truncated) Poisson distribution with mean parameter *m_t_*. We use the following values for *m_t_* to incorporate time-varying social distancing measures. The model was simulated using the Numpy library [39] in Python 3.

**Table 8:** The social distancing parameters for Model I.

| Time Range (in days) | $m_t$ |
| --- | --- |
| 1–7 | 5 |
| 8–14 | 4 |
| 15–126 | 3 |
| 127–210 | 4 |
| 211–217 | 5 |
| 217–366 | 6 |

#### C.2 Model II: A Stochastic SIR Model with Migration

For this model we consider a complete binary tree of depth k as in Figure 9 where *k* = 3. Such a tree has 2*^k^* leaves and *n* = 2*^k^*^+1^ – 1 nodes (including the leaves). There are *N* = 2*^k^* individuals and each of them is associated with a unique leaf. Every individual has a unique path between their leaf and the root, where the path consists of *k* +1 nodes (including the leaf and the root). At any point of time, every individual is located at one of the nodes on the path between its unique leaf and the root. A consequence is that individuals may be isolated with certainty in their leaf, or alternatively may be in the root or one of the other nodes of the tree where there is a possibility for them to be in physical proximity with other individuals. Thus the tree structure provides a spatial component (individuals are located at nodes) as well as a social component (individuals can meet specific groups of individuals only in specific parts of the tree).

**Figure 9:**
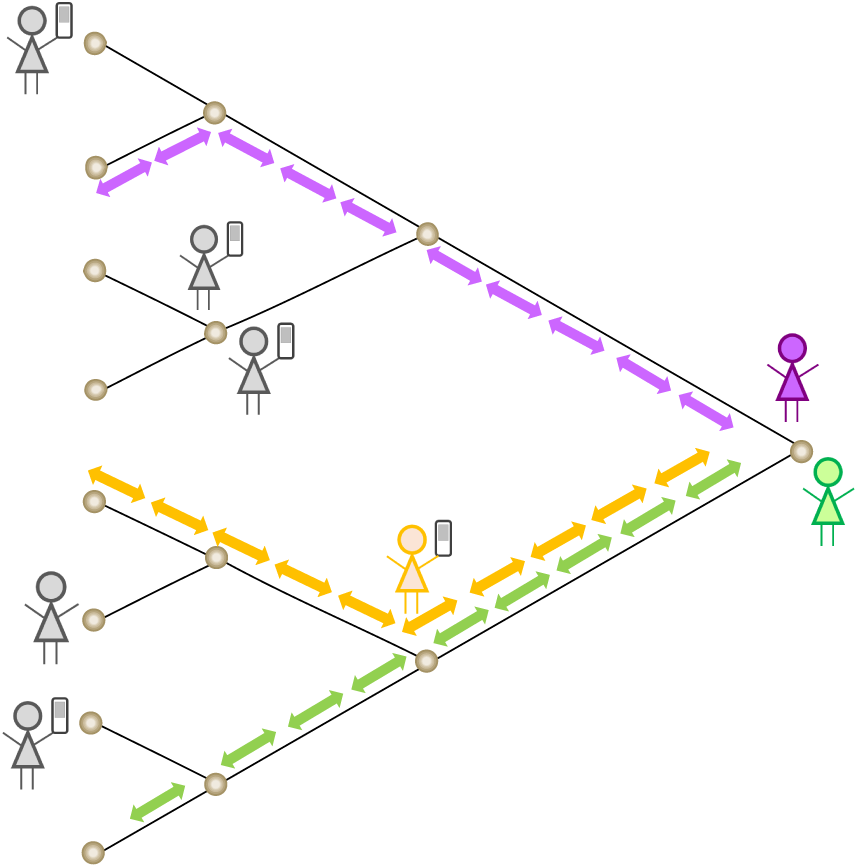
All *N* = 2*^k^* individuals traverse a binary tree between their private leaf and the root. At any node infection follows a continuous-time stochastic SIR model between the individuals present.

We say that the root is at distance k and the leaves are at distance 0. The movement of individuals occurs in continuous time and is in unit steps, either increasing distance by 1 or decreasing it by 1. Then for any distance *i* = 0,…, *k*, there is possibility to have up to 2*^i^* individuals in the node. Hence, the farther away (towards the root) that an individual travels, the larger the probability of having other individuals in physical proximity. This setup naturally yields a social distancing mechanism: we may enforce the individuals spend (on average) more time close to their leaves.

Both individual mobility and the epidemic dynamics (including Safe Blues dynamics) are governed by continuous-time Markov Chains. See, for example, [40] for an introduction to the continuous-time Markov Chain SIR model. The mobility of each individual along the leaf-root path follows a birth-death process on *i* = 0,…, *k*, with all of the 2*^k^* birth-death processes being independent. The birth rate is λ (constant for each distance level) and the death rates are *µ_i_* = *µi* (linearly increasing with the proximity to the root). Social distancing is enforced by increasing *µ*, which causes individuals to spend more time near or at their leaf nodes.

Individuals’ health state is subject to change via the standard SIR dynamics at each node in the tree. Specifically, if (at given node at a given time) there are *ℓ* individuals of which *ℓ_S_* are susceptible, *ℓ_I_* are infected and *ℓ_R_* are removed (with *ℓ* = *ℓ_S_* + *ℓ_I_* + *ℓ_R_*), then the rate of infecting other individuals at that node is *β_C_ℓ_S_ℓ_I_* (with the subscript *C* standing for COVID-19). Further, the rate of transitions from having *ℓ_I_* to *ℓ_I_* – 1 is *γC*ℓ_I_. Upon infection (removal), a random susceptible (infected) individual present at the node is selected for infection (removal).

In a similar manner to the COVID-19 dynamics, the individuals with Safe Blues enabled devices are subject to SIR dynamics for Safe Blues strands. Each Safe Blues strand is indexed by a unique integer *s*. The infection rate for strand *s* is *β_s_* and the corresponding removal rate is *γ_s_*. The dynamics are similar to the COVID-19 dynamics described above, except that at a given node only individuals with Safe Blues enabled devices take part.

##### Parameters used for the simulation run

We used a standard Doob-Gillespie simulation algorithm (see [41, Chapter 10]) to simulate the (time-varying) continuous-time Markov Chain of this model on the time range [0, 366]. This was for the case of *N* = 2,048 individuals (*k* = 11). The penetration proportion was *η* = 0.5 and thus *N^B^* = 1, 024.

The infection rate of COVID-19 was *β_C_* = 0.015 and the removal rate was *γ_C_* = 0.1. We simulated 10 Safe Blues Strands each with *γ_s_* =0.1 and *β_s_*= *U_s_*,*β_C_*/*η* where *U_s_* were pre generated i.i.d. uniform random variables on the range [0.5,1.5]. The initial infection proportion of both COVID-19 and Safe Blues strands was at 0.03.

For mobility within the tree we used λ = 0.9 and *µ_t_* was a time-varying rate as specified in Table 9.

**Table 9:** The social distancing parameters for Model II.

| Time Range | $\mu_t$ |
| --- | --- |
| [0,20) | 0.9 |
| [20,50) | 1.5 |
| [50,100) | 1.1 |
| [100,120) | 2.1 |
| [120,200) | 0.9 |
| [200,220) | 2.2 |
| [220,300) | 0.8 |
| [300,366] | 1.4 |

#### C.3 Model III: A Spatial Agent Model with Centrality

This model is based on N individuals moving on the Euclidian plane and it captures both spatial and social aspects of the interactions between individuals. If two individuals are in close proximity and one of them is infected with COVID-19, it can be transmitted to the other individual with a certain infection probability. The Safe Blue strands are transmitted in a similar way.

For each individual there is a unique fixed *base* (home), located at a fixed point on the plane. There are also commercial/social *centers* that attract individuals, also located at fixed points on the plane. Individuals can visit these centers each day independent of others. Individuals can have social interactions with neighbours near their homes, or at the centers during their visit. The spatial movement of each individual is captured using biased random walks.

The model evolves over time units of days, yet within each day there are finer small discrete time units in which individuals make small steps on the plane, always gravitating towards a fixed point which is either their *base* or a *center*. This gravitation is modelled using a biased random walk which is described later. In addition to the small discrete time steps, individuals may also make quick (immediate) transitions swapping their gravitational point of attraction from *base* to a *center* and vise versa. Whenever swapping occurs the new location around the destination (*base* or *center*) is chosen as a random point near the destination. This models quick transport (e.g. driving) between home and commercial/social centers. See Figure 10 for an illustration of the model.

**Figure 10:**
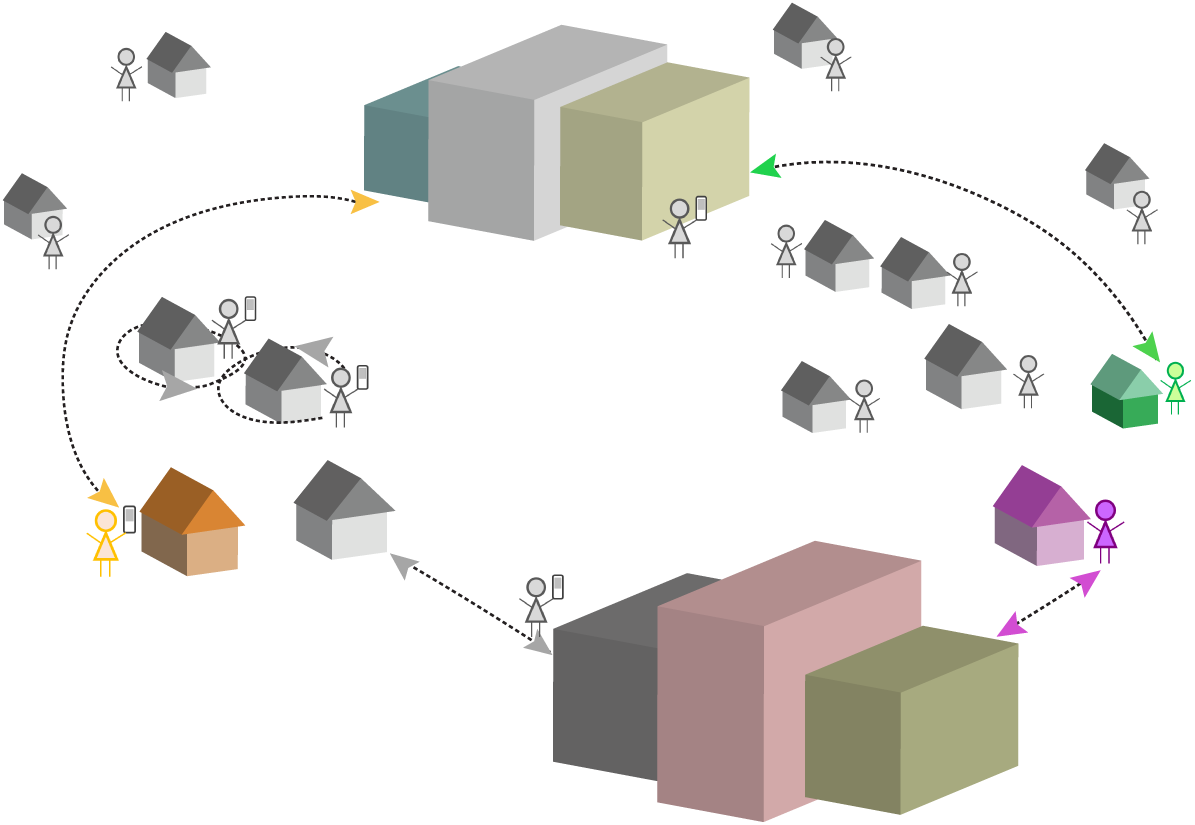
A spatial model where each individual diffuses either around their base or around a center.

The biased random walk that models the gravitation of individuals towards their unique *base* or towards a *center* depending if they are currently marked as “being at base”, or “being at a center” is executed by taking steps in a direction as follows. For individual *x*, consider the angle, *θ_x_*, between the individual and the attraction point (*base* or *center*). Then for some fixed parameter *κ*, we generate a random angle on [–π, π], following the Von Mises distribution with density

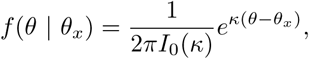

where *I*_0_(*κ*) is the modified Bessel function of order 0. Then a step with an exponential distribution having a small step size with time dependent mean *δ_t_*, is taken in the direction specified by the random angle.

The switching of the gravitational center is done as follows. On each day *t*, an individual spends time around (gravitating towards) the *center* during a time frame of length *w_t_*, which is selected uniformly and independently over the day. During the remaining time of the day the individuals spend time around (gravitating towards) their base. The choice of which center to move to is randomly selected proportionally to the Euclidean distance between the person’s current location and the center’s location. Hence people generally move to the center closest to their base, but not always.

Social distancing is enforced by modifying *w_t_* and *δ_t_* over time. When *w_t_* is low, individuals spend more time near their base and are less likely to meet others, while with *w_t_* large, individuals spend more time at centers and more social interaction is likely to occur. Further when social distancing is reduced (or increased) *w_t_* we also reduce (or increase) *δ_t_* for individuals currently at base. This implies that when social distancing is enforced, individuals are closer to home and when social distancing is relaxed, more interaction occurs.

In each time step during which an infected individual has another individual with a proximity of less than *r* distance units, the other individual may be infected with probability *p_C_* for COVID-19 and probability *p_s_* for Safe Blues strand *s*. On each day, the probability of removing an infected individual is *γ_C_* for COVID-19 and *γ_s_* for Safe Blues strand *s* similarly to the previous models.

At the onset of the simulation, the base locations are selected randomly and are fixed for the duration of the simulation. Further centers have fixed locations.

##### Parameters used for the simulation run

The simulation run that we created for experimentation had *N* = 5,000 individuals of which *N^B^* = 1, 000 had Safe Blues enabled devices (hence *η* = 0.2). The locations of the bases were generated at onset using a mixture of two bivariate normal distributions with means (25, 0) and (0, 0). The respective covariance matrices were,

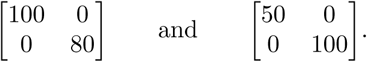

The mixture weighting between the distributions is at 0.3 and 0.7 respectively. There were 2 centers located at (10, 15) and (10, −15).

The simulation was run for 366 days where within each day there were 14 basic time steps. The parameter of the Von Misses distribution was set at *κ* = 6. The basic step size mean when not under social distancing was at *δ_t_* = 2. The proximity radius is *r* = 0.0085. The COVID-19 infection probability was set at *p_C_* = 0.04 and the recovery/removal probability was at *γ_C_* = 0.1/14. At onset a proportion of 0.0125 of the population was infected (uniformly initialized). Further, when individuals swap between a *center* and their *base*, they are located with a uniform angle around the destination and a distance that is exponentially distributed with mean 0.1.

For Safe Blues there were 50 strands denoted *s* = 1,…, 50, all with the same parameters. The strands were released at onset (*t*_start_ = 0). The removal probability was taken to be identical to COVID-19, and the infection probability was taken to be *p_s_* = *p_C_*/*η*. This rule follows the general guideline in Equation (1).

The parameters affecting social distancing, *w_t_* and *δ_t_* (used for base users), are expressed as functions of lockdown strength *l_t_* that takes values in the interval [0,1], where *l_t_* =0 indicates no lockdown and *l_t_* = 1 indicates highest possible lockdown. The details are summarized in Table 10 below. The model was simulated using the Numpy library [39] in Python 3.

**Table 10:**
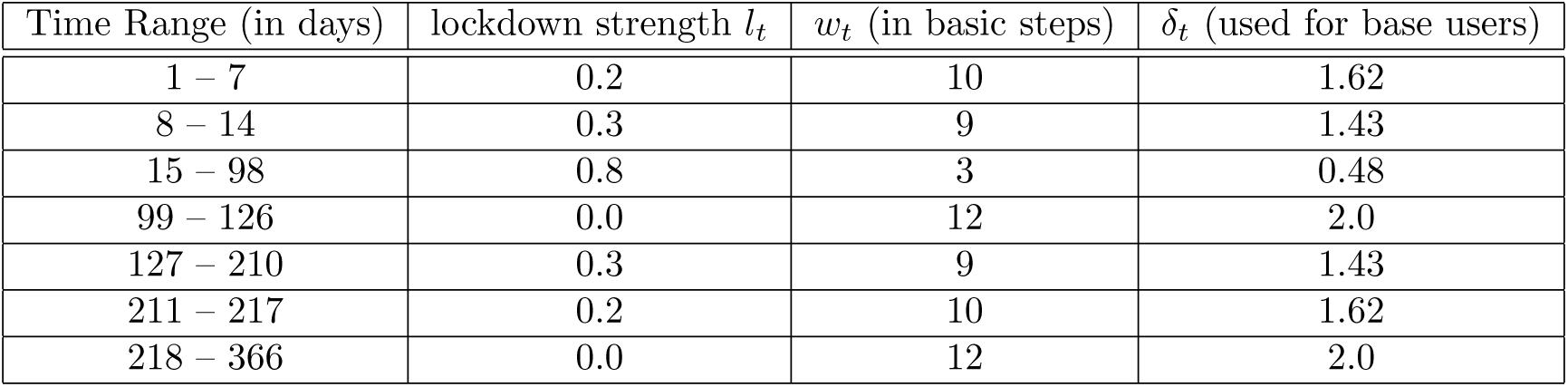
The social distancing parameters for Model III as functions of the lockdown strength, *l_t_*, defined by 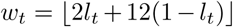 and *δ_t_* = 0.1*l_t_* + 2(1 – *l_t_*) (used only for base users), where 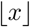 denotes the integer part of *x*.

## Notes

### Competing Interest Statement

The authors have declared no competing interest.

